# Small nucleolar RNAs in plasma and their discriminatory power as diagnostic biomarkers of Alzheimer’s disease

**DOI:** 10.1101/2021.05.04.21256076

**Authors:** Nicholas F. Fitz, Kyong Nyon Nam, Jiebiao Wang, M. Ilyas Kamboh, Radosveta Koldamova, Iliya Lefterov

**Author notes:** Correspondence to: Radosveta Koldamova, MD, PhD, Iliya Lefterov, MD, PhD.

## Abstract

The clinical diagnosis of Alzheimer’s disease, at its early stage, remains a difficult task. Advanced imaging technologies and laboratory assays to detect Aβ peptides Aβ42 and Aβ40, total and phosphorylated tau in CSF provide a set of biomarkers of developing AD brain pathology and facilitate the diagnostic process. The search for biofluid biomarkers, other than in CSF, and the development of biomarker assays have accelerated significantly and now represent the fastest-growing field in AD research. The goal of this study was to determine the differential enrichment of noncoding RNAs (ncRNAs) in plasma-derived extracellular vesicles (EV) of AD patients and Cognitively Normal controls (NC). Using RNA-seq, we profiled four significant classes of ncRNAs: miRNAs, snoRNAs, tRNAs, and piRNAs. We report a significant enrichment of SNORDs – a group of snoRNAs, in AD samples compared to NC. To verify the differential enrichment of two clusters of SNORDs – SNORD115 and SNORD116, localized on human chromosome 15q11-q13, we used plasma samples of an independent group of AD patients and NC. We applied ddPCR technique and identified SNORD115 and SNORD116 with a high discriminatory power to differentiate AD samples from NC. The results of our study present evidence that AD is associated with changes in the enrichment of SNORDs, transcribed from imprinted genomic loci, in plasma EV and provide a rationale to further explore the validity of those SNORDs as plasma biomarkers of AD.

## Introduction

Alzheimer’s disease (AD) is the most common and economically impactful form of dementia with no treatment. However, not only the exact clinical diagnosis of AD, at its early stage, is still a complex and uncertain task, but alongside the lack of efficient treatment, there are no established and reliable diagnostic tests to accurately identify patients who would benefit from newly developed therapeutic agents. Current criteria for AD diagnosis require well-defined clinical phenotype and pathophysiological biomarkers consistent with AD pathology (Dubois et al., 2014). An autopsy, however, remains the gold standard for diagnosing definite AD and, similarly to more than 115 years ago – microscopic Identification of amyloid plaques and neurofibrillary tangles in patients who meet the clinical criteria for probable AD. Importantly, detection of AD at the early stages of the disease has only become possible thanks to the development of biomarkers. The diagnostic guidelines published in 2011 recognize three imaging tests/neuroimaging biomarkers - amyloid PET, PET glucose uptake, and MRI (Budelier and Bateman, 2020). Diagnostic/biomarker tests using CSF, measure the levels of amyloid-beta (Aβ) peptides – Aβ_42_ and Aβ_40,_ total tau and phospho-tau (p-tau) (Milà-Alomà et al., 2019). While CSF biomarkers are widely used in research studies, currently, their clinical utility is considered appropriate only in select clinical situations – in the setting of a dementia clinic, in patients with early-onset, progressive or unexplained MCI or with comorbidities that add uncertainty to the diagnosis of AD (Shaw et al., 2018). The prohibitive cost of imaging tests and the invasiveness of the lumbar puncture, however, make the available tests impractical for enrolling and monitoring large number of asymptomatic participants in large clinical trials. Blood test to measure Aβ peptides, total and phospho-tau are much more appropriate tools, and regardless of their own and significant challenges, they have the potential for screening asymptomatic individuals and to aid in diagnosing symptomatic patients (Ashton et al., 2021). In evaluating the diagnostic accuracy of blood biomarkers, amyloid-PET or clinical diagnosis are considered reference (although hardly gold) standards (Budelier and Bateman, 2020). Since those two reference standards are imperfect, or “copper” standards, the reported calculated Sensitivity of the index test(s) can be raised or lowered (Kohn et al., 2013). Amyloid-PET, and all CSF and blood tests discussed above are specifically associated with APP processing, deposition of amyloid plaques and abnormal intracellular tau metabolism.

During the last 15 years or so, alongside the rapidly developing field of research of extracellular vesicles (EV), EV isolated from plasma of AD patients emerged as a novel and powerful potential biomarker of AD progression. The enormous potential of EV as diagnostic and treatment tools has made EV research in AD extremely attractive and highly significant (Dubois et al., 2014; Rajendran et al., 2014). EV are bilayer membranous structures that provide protection of their cargo from degradation and are produced by many cell types. Based on their mode of release and size, EV are divided into three classes: apoptotic bodies (500nm to 2μm) (Xu et al., 2019); microvesicles (also known as microparticles or ectosomes) (50nm to 1μm) (Chen et al., 2018), and exosomes (50nm to 150nm) (Jan et al., 2017). Because of their specific content and relatively easy access, EV have been at the center of intense research towards the identification of molecular biomarkers for diagnosis, prognosis and assessment of therapies in numerous pathological states, including AD (Lee et al., 2019; Paolicelli et al., 2019). Importantly, the content of EV cargo and membranes allows to differentiate and identify organ specificity-, including brain-specific EV (Rajendran et al., 2006; Chivet et al., 2012). Notably EV, regardless of their origin, are enriched with extracellular RNAs (exRNAs) of many different types, particularly noncoding RNAs (ncRNAs). The class of micro RNAs (miRNAs) has gained particular attention during the last 15 years, following the discovery of their export into the extracellular space in vesicles (Valadi et al., 2007) and quickly after that their release in bodily fluids (Chim et al., 2008). Thus, the idea that miRNAs can be used as molecular biomarkers to help diagnosing and monitoring a variety of pathological states was born and research to establish miRNAs as diagnostic biomarkers since attracted and utilized considerable funding. While there is no definitive conclusion if a specific miRNA or set of miRNAs exist for certain pathological condition(s), circulating miRNA research has not resulted in any highly specific, validated disease markers (Witwer, 2015; Tosar et al., 2021).

The goal of our study was to profile exRNAs in EV isolated from plasma of age-matched, early disease stage AD patients and cognitively normal control (NC) individuals. We identified ncRNAs in EV isolated from plasma of AD patients – carriers of *APOE4* allele *(APOE4+)* and *APOE3/3* genotypes and corresponding NC, and revealed differential enrichment of members of select classes of ncRNAs. We report for the first time a significant enrichment of SNORDs – C/D box small nucleolar RNAs (snoRNAs) in plasma EV of early-stage AD patients compared to NC. We discuss their applicability as diagnostic AD plasma biomarkers.

## Methods

### Study Population

#### Discovery Phase

Peripheral blood was collected from 39 individuals (21 females, 18 males) recruited by the Alzheimer’s Disease Research Center (PITT ADRC) at the University of Pittsburgh (Pittsburgh, PA) between 2015 and 2018 (Table 1). At inclusion the evaluation consisted of a Mini Mental State Examination (MMSE) and a Clinical Dementia Rating (CDR) tests. Withdrawal of blood and separation of plasma was also performed at inclusion. The cohort was divided into two main groups: a control group of 16 subjects with an MMSE score of 27 and above and a CDR of 0 considered cognitively normal (non-demented controls, NC); 23 patients with an MMSE under 27 and a CDR score between 0.5 and 2 were considered an AD group. After genetic testing for *APOE* genotype, the groups were further subdivided: of the 16 controls, 8 were *APOEε3/3* and 8 were *APOEε4/+*; of the 23 AD patients, 11 were *APOEε3/3* and 12 were *APOEε4/+*. Blood was collected in BD vacutainers with spray coated K2EDTA and centrifuged for 10 minutes at 2500g/4°C to eliminate cellular components. Plasma samples were stored at -80°C until further processing for EV isolation. **Verification Phase:** plasma samples from an independent group of AD patients and NC were used for verification. Twenty-four (24) individuals – 12 AD patients and 12 NC, were evaluated at PITT ADRC as described for those recruited in the validation phase (Table 2). The study was approved by the Institutional Review Board of the University of Pittsburgh.

**Table 1:** Discovery phase – patients’ demographics.

|  | NC (N=16) | NC-E3/3 | NC-E4+ | AD (N=23) | AD-E3/3 | AD-E4+ |
| --- | --- | --- | --- | --- | --- | --- |
| Female \| Male | 9 \| 7 | 5 \| 3 | 4 \| 4 | 12 \| 11 | 6 \| 5 | 6 \| 6 |
| Age<sup>a</sup> | 73 (6) | 73 (7) | 72 (4) | 76 (6) | 78 (6) | 74 (6) |
| Educaion<sup>b</sup> | 16.3 (2.6) | 17.0 (2.4) | 15.5 (2.8) | 15.2 (3.5) | 15.9 (3.9) | 14.6 (3.3) |
| MMSE<sup>c</sup> | 28.8 (1.0) | 28.8 (1.0) | 28.8 (1.0) | 20.1 (5.9) | 23.0 (3.1) | 17.9 (6.6) |
| CDR<sup>d</sup> | 0 | 0 | 0 | 0.83 (0.44) | 0.65 (0.24) | 0.96 (0.52) |

<sup>a</sup>, current age; <sup>b</sup>, years of education; <sup>c</sup>, last MMSE, and <sup>d</sup>, Last CDR. Data are mean (SD). NS by one-way ANOVA.

**Table 2:** Verification phase – patients’ demographics.

|  | NC (N=12) | NC-E3/3 | NC-E4+ | AD (N=12) | AD-E3/3 | AD-E4+ |
| --- | --- | --- | --- | --- | --- | --- |
| Female \| Male | 11 \| 1 | 6 \| 0 | 5 \| 1 | 8 \| 4 | 5 \| 1 | 3 \| 3 |
| Age<sup>a</sup> | 75 (6) | 74 (5) | 75 (8) | 79 (8) | 82 (3) | 76 (6) |
| Educaion<sup>b</sup> | 16.3 (2.7) | 16.0 (2.8) | 16.5 (2.8) | 13.5 (1.8) | 13.2 (1.6) | 13.8 (2.0) |
| MMSE<sup>c</sup> | 28.8 (1.3) | 29.2 (0.8) | 28.5 (1.6) | 19.3 (5.5) | 21.8 (6.3) | 16.7 (3.3) |
| CDR<sup>d</sup> | 0 | 0 | 0 | 1.08 (0.60) | 1.0 (0.55) | 1.17 (0.68) |

<sup>a</sup>, current age; <sup>b</sup>, years of education; <sup>c</sup>, last MMS, and <sup>d</sup>, Last CDR. Data are mean (SD). NS by one-way ANOVA; NC vs AD t-test p=0.059

### AD brain samples

All brain samples were provided for a previous study (Lefterov et al., 2019) by the University of Pittsburgh ADRC brain bank and the Sanders-Brown Center on Aging at the University of Kentucky, Lexington, KY. Brain tissue was not used or processed for this study. Here we only refer to the sequencing data, made publicly available at NCBI GSE144254.

### Isolation and sizing of EV

Plasma samples (500 µl each) were thawed on ice, diluted with 2 ml of cold PBS, press-filtered using a 0.8 μm filter, centrifuged for 20 min at 13,200 g/4°C and finally press-filtered again through a 0.22 μm filter. Using cleared plasma samples, EV were isolated by a two-step differential ultracentrifugation (dUC) \ procedure, each 70 min at 100,000 g/40C (Beckman Optima LE80K ultracentrifuge) was followed by final resuspension of the pellets in 50 µl of PBS. EV samples were stored at -80°C until further processing for EV characterization and RNA isolation. For EV measurements, after thawing on ice, 10 µL of the 50 µL EV suspension in PBS were diluted 1:100 in cold PBS to obtain 1 mL suspension. 660 µL of QIAzol (Qiagen) was added to the rest of the EV suspension and stored at -80°C until RNA extraction. For EV characterization, using precision electronic pump, 1 ml of diluted EV sample was loaded on a NS300 Nanosight instrument (Malvern Instruments) at a rate of 400 µL/min. The instrument was calibrated before each batch of samples using 2 µL of 100 nm latex beads diluted in 1 mL of PBS. Before and after the analysis of each sample, negative controls of PBS only were loaded and analyzed. Each sample analysis was composed of 5 separate measurements of 60 sec each. Each analysis of 5 separate 60 sec measurements were visualized and measured using Nanoparticle Tracking Analysis (NTA) software, version 3.4. EV size and concentration were compared by unpaired two-sample t-test (GraphPad Prism v8.4.2). Plasma EV were further characterized through detection of Flotillin, heat shock protein 70 (HSP70), CD63 and tumor susceptibility gene 101 (TSG101) by Western blotting and Transmission electron microscopy.

### RNA extraction and quality control

After thawing on ice, the 700 μl of EV suspension in QIAzol were processed to extract total RNA using MicroRNeasy micro kit (Qiagen) following the manufacturer’s instructions. Total RNA was purified using RNeasy MiniElute spin columns (Qiagen) and eluted with 15 µL of RNase-free water. Concentrations of RNA were determined using a QuBit (ThermoFisher), and quality was analyzed on a Bioanalyzer 2100 (Agilent).

### Library preparation and sequencing

To generate libraries, 1 ng of purified total RNA was fragmented and used to perform a first strand cDNA synthesis. RNA fragments were linked at the 3’ and 5’ ends to adapters and then to unique molecular identifiers (-UMI). Each sample received a unique combination of specific index/UMI during library generation. After conversion into cDNA using QIAseq miRNA Library Kit (Qiagen), the cDNA was finally purified for size selection and removal of short library fragments. The libraries’ quality and quantity were assessed on Bioanalyzer 2100 High Sensitivity DNA chip. Before sequencing, library sizes were evaluated using the High Sensitivity NGS Fragment Analyzer Kit (AATI) and quantified by qPCR using the KAPA qPCR quantification kit (KAPA Biosystems/Roche). Libraries were normalized and pooled as per manufacturer protocol (Illumina). Sequencing was then performed using the NovaSeq 6000 platform (Illumina) with 76 bp single-end reads on an SP-100 cycles flow cell.

### Computational analysis of sequencing data

Sequencing data were submitted to Genboree for pre-processing, alignment and mapping using the exceRpt pipeline (http://genboree.org)(Murillo et al., 2019; Rozowsky et al., 2019). Reads were aligned to human genome version hg38 (https://genome.ucsc.edu/) and assigned as miRNA (miRBase), tRNA (gtRNAdb), piRNA (piRNABank), longRNA (GENCODE), circRNA (circBase) or snoRNA (GENCODE). The remaining reads were mapped to exogenous miRNAs in miRBase and rRNAs in the Ribosomal Database Project (RDP). Read counts were then used for further analysis. To identify and assess differences in enrichment of ncRNAs in plasma EV, we performed differential expression analysis using DESeq2 and Wald test, as implemented in Genboree. We applied a two-step RT-qPCR (TaqMan microRNA assays and TaqMan Noncoding RNA Assay, ThermoFisher), using RNA isolated for sequencing, on a ViiA 7 Real Time PCR System (Applied Biosystems) to verify sequencing results of selected miRNAs and snoRNAs.

### Droplet digital PCR

In the verification phase, we performed absolute quantification of expression level of snoRNAs/SNORDs extracted from plasma EV by droplet digital PCR (ddPCR). Total RNA was reverse transcribed using iScript Advanced cDNA Synthesis Kit for RT-qPCR (BioRad). The PCR reaction was performed in oil droplets using the ddPCR supermix (BioRad) and TaqMan non-coding RNA assays (ThermoFisher), according to manufacturer’s suggested protocol (BioRad). Analysis of oil droplets was conducted by QX200 Droplet Reader and absolute number of targeted SNORD copies were determined and analyzed by QuantaSoft Software v1.7 (BioRad). Each assay included a negative control for each RT reaction and a no-template control according to the protocol provided by BioRad. Significance was determined by unpaired two-sample t-test (GraphPad Prism v8.4.2).

### Statistics

Subjects’ age was analyzed by unpaired two-sample t-test and one-way ANOVA. EV size and concentration and PCR data were analyzed by unpaired two-sample t-test and one-way ANOVA followed by Tukey’s post hoc multiple comparisons test. Plasma transcriptome data were analyzed by Wald test using DESeq2 as implemented in Genboree (http://genboree.org/site/), and postmortem brain transcriptome data were analyzed by quasi-likelihood F test using edgeR. ncRNAs were considered significantly differentially enriched when p<0.05. All t-tests or ANOVAs were performed using Microsoft® Excel for Mac, version 16.33 and GraphPad Prism, v8.4.2.

## Results

### Identification and differential analysis of enriched ncRNAs in plasma EV of AD patients and NC

We isolated and characterized EV from plasma of AD patients and NC. All 39 samples – APOE3/3 and APOE 4/+ genotypes, were randomly chosen from a series collected routinely at ADRC Pittsburgh between 2015-2018 (Table 1). AD was classified based on CDR and MMSE scores. Individuals with an MMSE scored 27 and above and CDR of 0 were NC. Those with an MMS scored 27 and below and CDR equal to or above 0.5 were considered AD.

Plasma EV were characterized based on size, concentration, and protein composition. The analysis of the Finite Track Length Adjustment (FTLA) for concentration and particle size revealed a significant enrichment of EV with an average size of 104.6 ± 2.8 nm and concentration of 6.22×10^7^ ± 1.08 particles/ml (Figure 1A). Since most of the detected particles were measured between 50nm and 200nm, this indicated an enrichment of EV with small vesicles that greatly reduced the relative amount of much lar ger EV, such as apoptotic bodies. No difference of EV size or concentration was noticed between AD and NC groups (Figure 1B and C). The enrichment of EV obtained by ultracentrifugation was confirmed by detecting four different EV markers by Western blot: Flotillin, heat shock protein 70 (HSP70), the CD63 antigen and tumor susceptibility gene 01 (TSG101, a component of the ESCRT-I complex, a vesicular trafficking process regulator) (Figure 1D) and verification of size by Transmission electron microscopy (Figure 1E).

**Figure 1:**
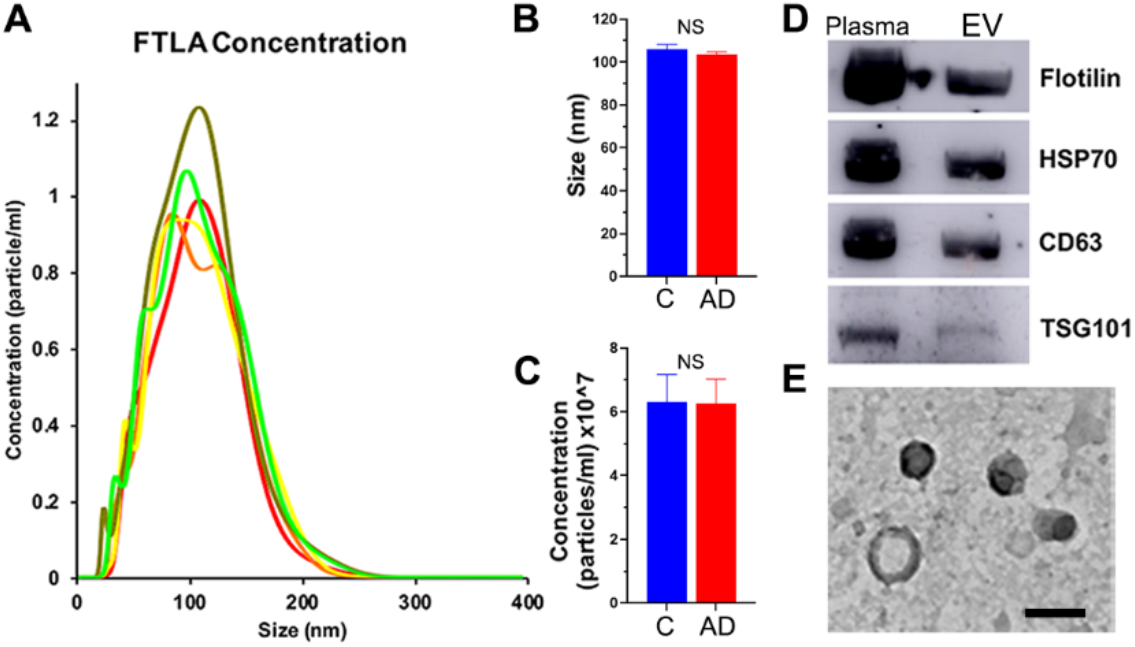
Characterization and quantification of isolated plasma EV. (**A**) Representative results of nanoparticle tracking analysis (NTA) after 5 successive measurements of diluted plasma EV. Average size (**B**) and concentration (**C**) of EV in AD and Control samples. Statistical significance was determined by unpaired t test. NS = No Significance. Western Blot of EV markers (**D**) and representative EM images (**C**) of EV confirming average

The RNA-seq libraries were evaluated for quality control and sequenced on Illumina NovaSeq 6000 System. The sequencing data were analyzed using the exceRpt pipeline as implemented in Genboree (Murillo et al., 2019; Rozowsky et al., 2019) and we determined differential expression between AD and NC. The analysis using exceRpt revealed the full composition of the ncRNA population across the 39 sample s. For the analysis we selected genes with average CPM > 5 (log10CPM > 0.7) and identified 512 genes that passed this cut-off (Supplemental Table 1). The majority of those were miRNAs (69.7%), followed by small nucleolar RNAs (snoRNAs) 21.6%, transfer RNAs (tRNAs) 4.7% and Piwi-interacting RNAs (piRNAs) 3.9% (Figure 2A-B). Among these 4 classes we identified 57 differentially affected transcripts when comparing AD vs NC: 34 snoRNAs (Figure 2C and F), 15 miRNAs (Figure 2D), 6 piRNAs (Figure 2E) and 2 tRNAs (Supplemental Table 1). To validate sequencing data, RT-qPCR verification was performed on select miRNAs and Box C/D snoRNAs (SNORDs) with total RNA isolated for sequencing and TaqMan expression assays (Supplemental Fig. 1).

**Figure 2.**
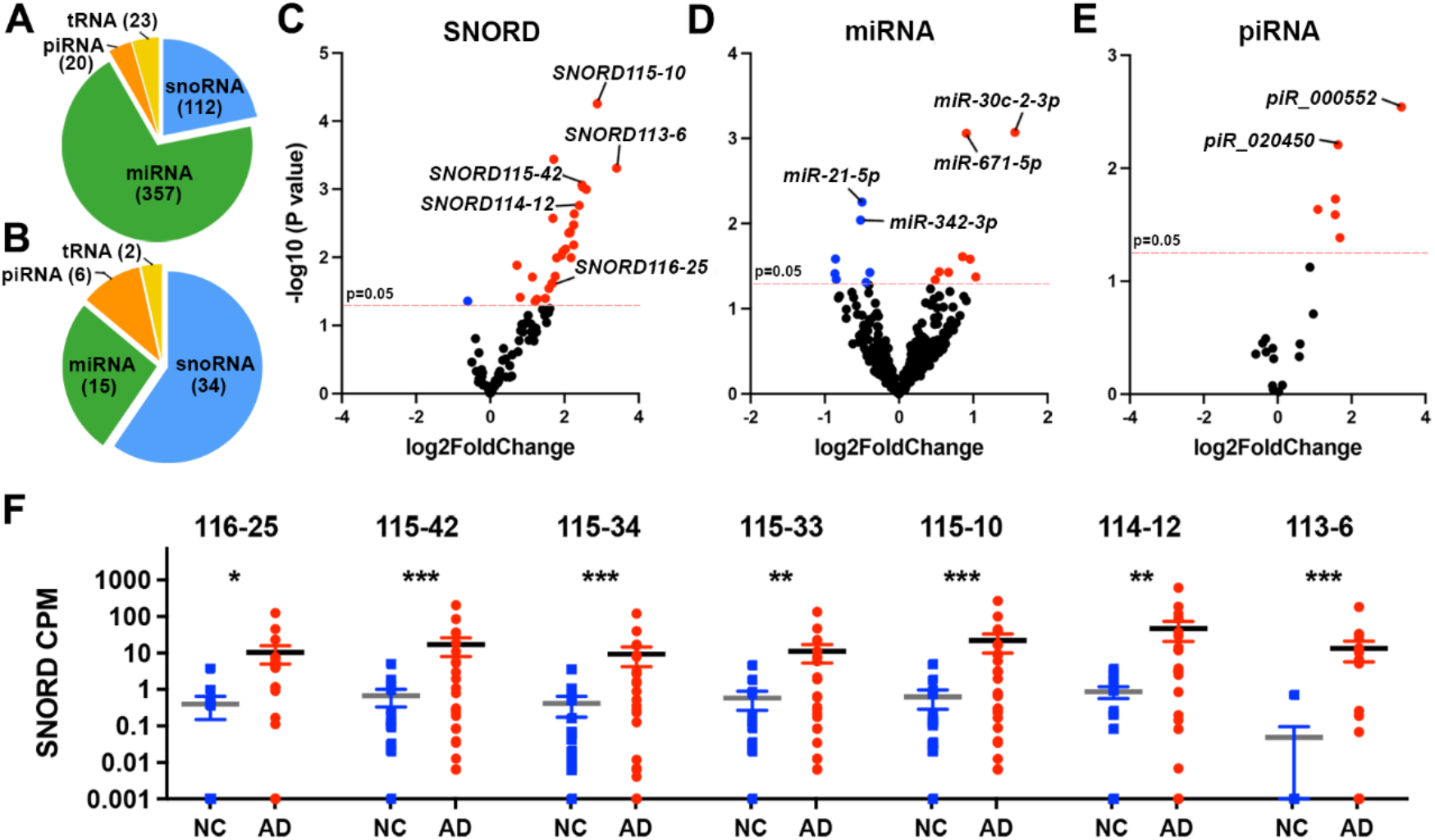
Enrichment of SNORDs associated with AD vs Cognitively normal controls (NC) in plasma EVs. (**A**) Distribution of non-coding RNAs identified in plasma EVs from all human samples in the Discovery phase at a cutoff of Log10CPM >0.7. (**B**) Distribution of four classes of differentially enriched non-coding RNAs in plasma EVs from AD patients compared to NC. (**C-E**) Volcano plots showing differential enrichment of SNORDs (**C**), miRNA (**D**) and piRNA (**E**). (**F**) Scatter dot plot of RNA-seq data; CPM of select SNORDs in EVs of AD vs NC. For C, D, E and F, the cutoff is Log10CPM>0.7 and p<0.05. Statistical significance was determined by exceRpt. Error bars represent mean ± SEM. N=15 (9 females), NC; N=24 (12 females), AD patients. *** p<0.001; ** p<0.01; * p<0.05.

Subsequent analysis was focused on SNORDs, based on their abundance in EV isolated from plasma samples and their reported expression in brain. In humans, the vast majority of sno RNAs are encoded primarily within introns of host genes, which by themselves may be noncoding (Dieci et al., 2009; Yang et al., 2019). Based on structural characteristics and associations with canonical partner ribonucleoproteins, snoRNAs coded by intronic host genes are classified into either C/D- (SNORD) or H/ACA-box (SNORA) subfamilies (Yu and Meier, 2014; Cavaillé, 2017; Bratkovič *et al*., 2018; Deogharia and Majumder, 2018; Bratkovič *et al*., 2019).

In plasma EV from AD vs NC, we identified differentially enriched SNORD clusters: SNORD113 (2 genes), SNORD114 (3 genes); SNORD115 (17 genes) and SNORD116 (3 genes). SNORD113/SNORD114 and SNORD115/SNORD116 are tandemly repeated C/D box snoRNA clusters of reciprocally imprinted genes residing in imprinting control regions (ICR) of the human 14q32.2 and 15q11-13 domains, expressed from paternally and maternally inherited alleles (Cavaille et al., 2000; Cavaillé et al., 2002; Kishore et al., 2010; Chamberlain, 2013; Cavaillé, 2017; Chung et al., 2020).To evaluate how significant the presence of the members of these four SNORD clusters in plasma EV is, we used the information available in snoDB database: (http://scottgroup.med.usherbrooke.ca/snoDB/), an online interactive database tool which provides consolidated information on snoRNAs, including interpretations of available high throughput sequencing data and organ specific expression (Bouchard-Bourelle et al., 2020).

In both genotypes (Supplemental Table 1) differentially enriched transcripts of SNORD115 cluster appeared with a higher abundance in AD samples compa red to NC (few genes are marked on Figure 2F). In humans, SNORD115 is among the very few snoRNAs with organ-specific expression: this cluster of non-coding RNAs is expressed exclusively in brain and specifically in neurons (Wevrick et al., 1994; Chamberlain and Lalande, 2010b; Bortolin-Cavaille and Cavaille, 2012; Chamberlain, 2013; Cavaillé, 2017; Chung et al., 2020; Keshavarz et al., 2020). As shown on Supplemental Fig. 2, we found a similar effect in AD patients of E3/3 and E4+ genotypes vs their respective controls. We identif ied transcripts of 2 SNORD113 gen es – SNORD113-6 and -9, and 3 g enes, and members of SNORD114 cluster – SNORD114-2, -3 and 12 differentially enriched in EV isolated from AD plasma samples. According to the sequencing data available in snoDB the members of SNORD113 and SNORD114 clusters identified differentially enriched in our study are relatively highly expressed in liver, breast, ovary and prostate(Dsouza et al., 2021).

### Verification of differentially enriched SNORDs in plasma EV of an independent group of AD patients and NC

We verified the validity of the results in an independent group of AD patients and NC (Table 2), with similar inclusion criteria as for the discovery group: MMSE and CDR scores, and confirmed APOE genotype. We chose SNORDs for verification that are exclusively/predominantly expressed in brain and highly enriched in EV of AD plasma – SNORD115 and 116; 2) highly enriched in EVs of AD plasma – members of SNORD113 and SNORD114 clusters. Using RNA extracted from plasma EV of 12 AD and 12 NC samples, we performed absolute quantification of enrichment levels of the above SNORDs by digital droplet PCR (ddPCR) and analyzed the results to test: a) the difference AD vs NC for each of the SNORDs; the goal was to reveal an association between significantly abundant SNORDs in plasma EV and the disease status, and b) to determine if there were SNORD predictors that can be useful and further tested as diagnostic plasma biomarkers for AD. As shown on Figure 3 A and C, for both SNORD115 and SNORD116 there was a significant difference between AD and control samples. Moreover, the difference was preserved when AD samples were compared to controls within each APOE genotype (Figure 3B and D). Importantly, the level of enrichment of SNORD115 was higher in AD-E4+ samples compared to AD-E3/3 (Figure 3B). The result, even considering the relatively small number of participants in the verification group, clearly indicated *APOE* allele associated effect. While members of clusters SNORD114 and SNORD113 were identified as differentially enriched by RNA-seq, those differences were not confirmed by ddPCR (data not shown).

**Figure 3:**
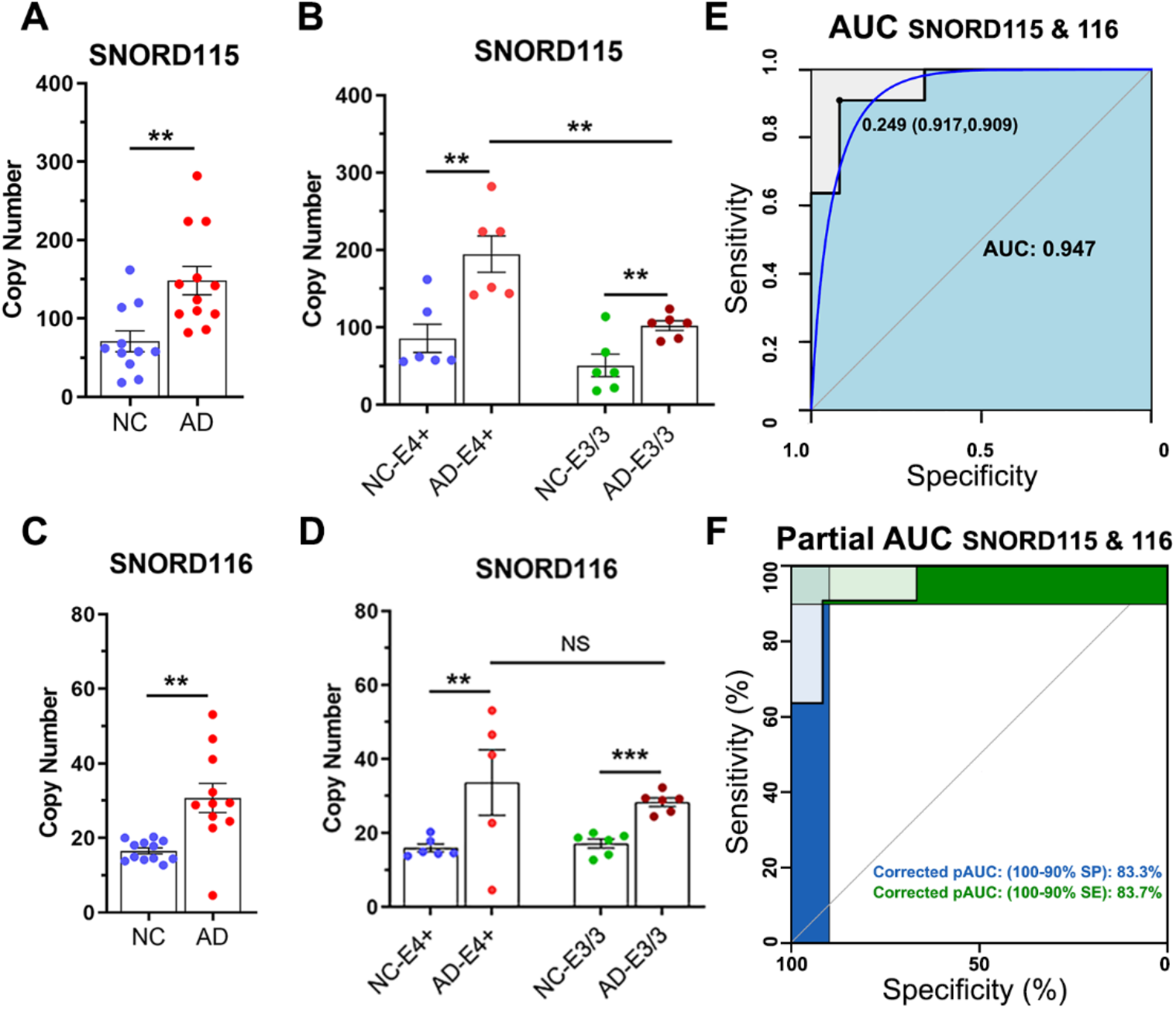
Enrichment of SNORDs in plasma EV as AD predictors. In the verification phase EV were isolated from plasma samples of a separate cohort of AD patients and NC individuals (independent of the Discovery phase) and assessed using ddPCR. (**A-B**) Scatter plots depict the abundance of SNORD115 (**A**) and 116 (**B**) in plasma EV isolated from NC (N=12) and AD patients (N=12). Statistical significance determined by unpaired *t*-test. **(C-D**) Scatter plots depict the effect of *APOE* genotype on enrichment of SNORD115 (**C**) and SNORD116 (**D**) in EV of NC and AD plasma samples. Statistical significance determined by *t*-test. ** p < 0.01; *** p < 0.001; NS, No Significance; N=6 per isoform. (**E-F**) ROC analysis showing the AUC (**E**) and corrected Partial AUC (**F**) for combined SNORD115 & 116 data. The blue line on **E** is the smoothed ROC curve. In addition to AUC we show the copy number threshold with the highest sum of sensitivity and specificity.

To identify AD predictors, Receiver Operating Characteristic (ROC) curves were calculated for SNORD115 and SNORD116; the data for those calculations were absolute copy numbers as determined for th e samples of the verification groups by ddPCR. A cutoff value was selected as the threshold predicting AD with Area Under the Curve (AUC) > 0.8. The values of AUC for SNORD115 – 0.861, as well as for SNORD116 – 0.886 exceeded the threshold, thus according to widely accepted classification within the range of “outstanding” (Mandrekar, 2010; Konstantinou et al., 2012). The value of combined AUC calculated for both, SNORD115 and SNORD116, was 0.947 (Figure 3 E,F)

### Differentially enriched SNORDs in brain of AD patients and NC

In a recently published study, we performed bulk RNA-seq using postmortem brain samples from posterior parietal lobule of AD patients of 3 APOE genotypes (APOE2/carriers, APOE3/3 and APOE4/carriers) (Lefterov et al., 2019). Here, we compared transcriptomic profiles of APOE3/3 vs APOE4+ (see demographics on Table 3) and identified 102 SNORD genes (Figure 4A, Supplemental Table 2). There were 12 SNORD transcripts differentially expressed between AD-E4+ and AD-E3/3 (Figure 4A and B). As shown on Figure 4C, we found that the expression level of several SNORDs hosted by the Small Nucleolar RNA Host Gene 14 (*SNHG14*) located on human Chr15q11-q13 was higher in AD patients of APOE4/+ genotype, compared to APOE3/3.

**Table 3:** Postmortem brain samples.

|  | AD-E3/3 (N=12) | AD-E4+ (N=22) |
| --- | --- | --- |
| Female \| Male | 7 \| 5 | 10 \| 12 |
| Age<sup>a</sup> | 85 (4) | 84 (4) |
| PMI (hrs) | 4.1 (2.2) | 4.3 (2.9) |
| Braak | 6 | 6 |

<sup>a</sup>, age at death; Data are mean (SD). NS by t-test

**Figure 4.**
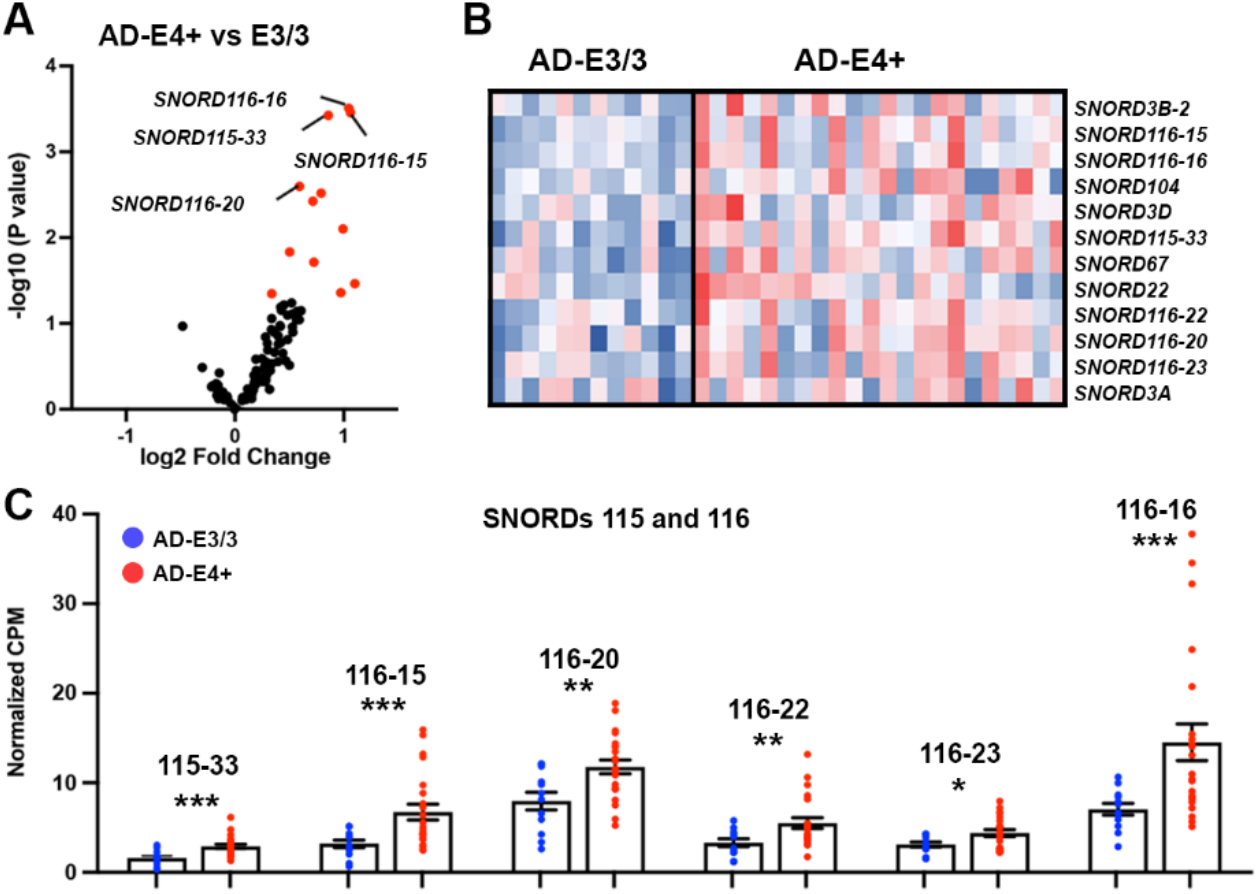
Increased expression levels of select SNORD115 and SNORD116 genes in E4+ vs E3/3 postmortem AD brains. RNA-seq analysis was performed on postmortem samples from posterior parietal lobule of E3/3 and E4+ AD patients. N=12 (7 females) AD-E3/3, N=22 AD-E4+ (10 females). Statistic is by edgeR. (**A**) Volcano plot shows the enrichment of SNORDs in brain of AD-E4+ vs AD-E3/3 postmortem brain samples. red dots denote the differentially expressed transcripts. (**B**) Heatmap shows 12 significantly up-regulated SNORDs in AD-E4+ vs AD-E3/3 at p<0.05. (**C**) Scatter dot plot depict CPM of statistically significant SNORD115 and SNORD116 genes from AD-E4 and AD-E3/3 postmortem brain samples: * p < 0.05; ** p < 0.01, ***

## Discussion

For more than 20 years now miRNAs have dominated the field of research on ncRNAs and post-transcriptional regulation of gene expression. Not surprisingly, the identification of miRNAs in EV, visualization of their transf er between cells and protected release into the bloodstream, immensely stimulated the field of biomarkers research. The idea is that distinct developmental and pathological processes are associated with transcription of unique miRNAs with highly important functional and regulatory relevance. For example, miRNAs have been implicated in AD through multiple pathways, including lipid metabolism, neuroinflammation, tau-phosphorylation and expression of amyloid precursor protein (APP) (Maoz et al., 2017). Moreover, once packed into EV and released into the bloodstream the detection of disease specific miRNAs in EV would facilitate the diagnosis of asymptomatic disease process at early stages, screening and monitoring the effect of newly developed therapeutic agents. This idea generated a significant informational bias towards miRNAs (Witwer, 2015; Maoz et al., 2017), which holds true for neurological disorders as well, including AD, even today.

Differential expression of snoRNAs has been observed in brain of AD model mice and were associated with the initial progression of amyloid depositions (Gstir et al., 2014). Differential expression of snoRNAs in dorsolateral prefrontal cortex of postmortem brains of patients with Autism Spectrum Disorder was reported more than 5 years ago (Gstir et al., 2014; Wright et al., 2017). We have recently reported a correlation of snoRNAs with *APOE* genotype in postmortem brain tissue of AD patients [(Lefterov et al., 2019); see also Figure 4]. Association of piRNAs with AD and neuronal activity (Wakisaka and Imai, 2019) or a suggestion for their use as CSF AD biomarkers in combination with miRNAs has been published (Jain et al., 2019; Wakisaka and Imai, 2019).

The goal of our study was to determine if EV isolated from plasma of AD patients and non-demented individuals contain ncRNAs differentially associated with the disease. We did not identify miRNAs expressed exclusively or even predominantly in brain and enriched in plasma EV samples from AD patients. To the best of our knowledge there have not been any reports so far that miRNAs exclusively expressed in brain exist, nor have there been miRNAs exclusively associated with AD, moreover contained in, and isolated from plasma EV. Despite the recent interest in piRNAs as biomarkers, especially in brain and dementia, little is known about their role in AD (Qiu et al., 2017; Roy et al., 2017; Jain et al., 2019; Mao et al., 2019). Most of the studies have been conducted on postmortem brain tissues and showed patterns of piRNAs associated with AD (Qiu et al., 2017; Roy et al., 2017; Mao et al., 2019). In their study performed on exosomes (authors’ definition) isolated from CSF (Jain et al., 2019), Jain et al. presented a signature of 3 miRNAs and 3 piRNAs, suggesting that they could be suitable as new biomarkers to detect AD. The fact that the identified plasma piRNAs in our study didn’t overlap with Jain et al. study performed on EV isolated from CSF (Jain et al., 2019) is not surprising. Studies based on CSF, plasma and whole blood have been shown to present very different ncRNA signatures, with some of them being up-regulated in CSF and down-regulated in blood from the same study (Yanfang Zhao, 2019). The results of our study indicate that piRNAs expressed in human brain (hsa_piR_001042 and hsa_piR_010894 (Roy et al., 2017)) if combined with other ncRNA could be of interest as potential new bioliquid biomarkers of AD.

While RNA modifications guided by snoRNAs have been known for about more than 70 years now (Cohn, 1959), neither the spectrum of their biological functions nor the biogenesis of other regulatory ncRNAs following the processing of snoRNA transcripts have been well understood (Kishore et al., 2010). We detected a significant number of snoRNAs differentially enriched in plasma EV of AD patients compared to NC. To the best of our knowledge, this is the first report that snoRNAs are differentially enriched in EV isolated from plasma of AD patients. SNORD115 and SNORD116 are clustered in intronic regions of an imprinted host gene and are known to be expressed predominantly – SNORD116, or exclusively – SNORD115, in brain (Chamberlain and Lalande, 2010a; Chamberlain, 2013; Lässer et al., 2016; Cavaillé, 2017; Driedonks et al., 2018; Irimie et al., 2018; Chung et al., 2020). These clusters are transcribed within the same *SNHG14/SNURF-SNRPN* locus. Therefore, the presence of numerous SNORD115 transcripts in EVs, identified as significantly enriched in AD plasma samples suggests that a significant number of expressing units within *SNHG14* are actively transcribed in all AD patients. In plasma EV of an independent cohort of AD patients and NC we verified the differential enrichment of SNORD115 and SNORD116 by ddPCR (Figure 3). ROC analysis revealed SNORD115 and SNORD116 with “excellent” values of AUC (> 0.8). Their combined discriminatory power – we cautionary call those “AD SNORD signature”; has a combined AUC calculated as 0.947, which is considered outstanding and, to the best of our knowledge, outperforms any of the ncRNA signatures published so far as plasma AD diagnostic biomarkers. The combined AUC calculated for p-tau and Aβ42/40 measured in CSF rarely reaches 0.9 (https://meetings.alzdiscovery.org/wp-content/uploads/2020/05/ADDF-conference-Iswariya-Venkataraman_EUROIMMUN.pdf). Moreover, there has been only one study reporting a combined AUC of 0.98 for p-tau and Aβ42/40 with calculations based on CSF ELISA assays, plus a signature of combined 3 miRNAs + 3 piRNAs with scores derived by deep sequencing of RNA isolated from CSF samples (Jain et al., 2019). We report values of individual AUCs for SNORD115 and SNORD116, measured in plasma EV by ddPCR, calculated as 0.86 and 0.88, respectively.

The Identification of members of SNORD115 and SNORD116 clusters significantly abundant in peripheral blood of AD patients raises questions. **First: Do the SNORDs identified in plasma EV come from brain?** The tissue specificity of plasma biomarkers is of utmost importance, regardless of the biofluid. In this regard, it is important to mention, that the canonical function of SNORDs is guidance of posttranscriptional 2’-O-methylation of ribosomal RNAs. *SNORD115* are *SNORD* genes (together with several other (Bratkovič *et al*., 2019) without anti-sense sequence complemen tarity (next to their D/D’ boxes) to canonic al targets - ribosomal RNAs (Chamberlain, 2013). Interestingly, these so called “orphan” SNORDs are expressed in a tissue-specific manner. All genes of SNORD115 cluster, for example are exclusively expressed in brain (Rogelj, 2006; Chamberlain, 2013; Jorjani et al., 2016). Although it has been reiterated in recent publications, (Cavaillé, 2017; Deogharia and Majumder, 2018; Chung et al., 2020), there is evidence that malignant cells of lung cancer tissue in primary cultures and lung cancer cell lines express members of SNORD115 cluster, in addition to many other SNORDs and SNORAs (Liao et al., 2010; Gao et al., 2014; Mourksi et al., 2020). Engineered expression of members of SNORD115 in brain tissues other than brain parenchyma – choroid plexus, has also been reported (Raabe et al., 2019). However, we have not been able to find a report that SNORD115 or individual members of SNORD115 cluster were released in plasma EV of patients with lung cancer (Steinbusch et al.; Nossent et al., 2019). Members of SNORD115 and SNORD116 clusters have been also reported upregulated in patients with multiple myeloma, chronic lymphocytic leukemia (Ronchetti et al., 2012; Ronchetti et al., 2013), or acute lymphoblastic leukemia (Vendramini et al., 2014). Within the list of all SNORDs associated with myeloma reported by Ronchetti et al. (Ronchetti et al., 2012), in our study we found only SNORD115-32 and SNORD116-29 overlapping with the SNORDs identified by RNA-seq in plasma EV. Importantly, for both SNORDs we calculated poor AUCs and therefore they did not justify further testing. While Ronchetti et al. (Ronchetti et al., 2012; Ronchetti et al., 2013) did not specifically look for ncRNAs in plasma or serum obtained from their patients, it is reasonable to assume that members of SNORD115 and SNORD116 clusters could be identified in EV, in RNA:proteins or RNA:HDL complexes released by malignant B or T cells. Since none of the AD patients and NC in our groups were diagnosed with lymp homa or leukemia, we assume that members of SNORD115 in EV of plasma samples presented in this study come from brain. Interestingly, in meningioma samples – members of SNORD115 cluster have not been identified as up-regulated, supporting their exclusive neuronal origin (Jha et al., 2015; Rynkeviciene et al., 2018). While highly specific neuronal expression of SNORD115 facilitates their rapid and efficient detection in biofluids by ddPCR, isolation of brain specific EV using immuno-affinity approaches should be considered in a near future. **Second**: **Does a differential abundance of SNORD115s in AD plasma EV reflect an impaired metabolic or regulatory pathway with a role in AD pathogenesis?** In brain, SNORD116 and SNORD115 clusters have been primarily associated with Prader-Willi syndrome (PWS) (Bortolin-Cavaille and Cavaille, 2012; Cassidy et al., 2012; Chamberlain, 2013; Cavaillé, 2017; Deogharia and Majumder, 2018; Baker et al., 2020; Chung et al., 2020). In the majority of PWS cases, there is a deletion of the entire *SNURF-SNRPN* locus on the paternal human chromosome 15q11-q13 where *SNORD115* is mapped (Nicholls et al., 1993; Cavaillé, 2017). However, fine mapping of the centromeric 15q11-q13 region showed that the complete loss of *SNORD115* was not associated with obvious clinical phenotype (Runte et al., 2001), and in a case of confirmed PWS, the minimal deletion did not include *SNORD115* cluster (Kishore et al., 2010). In a recent article, J. Cavaillé group using CRISPR/Cas9-mediated *Snord115* knockout mouse raised questions concerning SNORD115 transcripts’ physiological role, notably the involvement of the cluster in behavioral disturbance associated with PWS (Hebras et al., 2020). Functionally, human *SNORD115* cluster has been associated with, and so far only, post-transcriptional modification of serotonin receptor subtype 5 (5-HT2C) mRNA (Cavaillé, 2017). Based on SNORD115 sequence complementary to *5-Ht2c* it was postulated that mouse SNORD115s regulate, or at least take part in the alternative splicing of *5-Ht2c* pre-mRNA. Surprisingly, a small effect of 5-HT2C in the development of only certain symptoms of AD has been suggested (Pritchard et al., 2008), but the involvement of 5-HT2C in the pathogenesis of AD has not been confirmed. Changes in function of 5-HT2C in AD is a topic beyond the scope of this study. It is worth mentioning, however, that the modulatory effect of SNORD115 on 5-HT2C expression is not mediated by the canonical enzymatic activity of Fibrillarin (Cavaillé, 2017; Deogharia and Majumder, 2018). To answer all those questions, future studies with larger groups of patients at different phases of the disease applying different “high specificity, high recovery” EV isolation procedures (Thery et al., 2001) and validation of the results in CSF and postmortem brains matching CSF and plasma samples are required. The lack of answers now, however, does not diminish the significance of SNORD115 transcripts (SNORD115/SNORD116, “AD SNORD signature”) in plasma EV and their potential role as diagnostic biomarkers of AD. **Third, but not least: is there APOE isoform dependent effect on the abundance of SNORDs in AD plasma EV and if yes, what is the mechanism**. One could imagine that APOE isoform dependent effect could be addressed, confirmed or ruled out, just by evaluating the levels of SNORD115 and/or the combination of SNORD115 & SNORD116 in plasma EV of sufficiently large group of age and sex matched AD patients and NC. This only looks like an easy task. Recruitment of NC of E4/4 genotype and AD-E2/2 patients is enormously difficult. The differential expression of SNORD115 and SNORD116 genes in postmortem brains of AD patients, however, even though without comparing the levels to postmortem brains of NC (see Figure 4) is a clear indication that a genotype effect is likely to exist. If this effect holds true in a much larger set of postmortem brains and other brain areas, in addition to posterior parietal, it will justify the hypothesis that the genotype effect materializes at a gene level, or through epigenetic mechanisms – histone modifications or changes in chromatin architecture. It will also justify resequencing of SNORD115 and SNORD116 clusters in easily accessible samples – PMBC, of a large population of NC and AD patients. The second part of the above question is also very difficult to answer. Outside the gene/gene interactions and epigenetic mechanisms, each of the possible APOE isoform dependent scenarios – facilitated packaging, fusion of multivesicular bodies, release of EV through the neuronal cell membrane and preferential clearance of EV from brain interstitial fluid through BBB and Blood-CSF-barriers, are beyond the initial goal of this study and will require substantial additional research. Altogether, however, without diminishing the significance of the APOE genotype effect, the disease effect, on the enrichment of SNORDs in AD plasma EV by itself, is a remarkable addition to the AD plasma biomarkers research and is worth further efforts.

There are **limitations** of this study which need to be taken into consideration. Because the differential ultracentrifugation with initial low speed centrifugation and filtering provides intermediate recovery and specificity of EV (Théry et al., 2018), we cannot exclude the small possibility of contamination of the pellet with vesicles smaller than 30nm, neither with those larger than 100 nm. Similarly, sin ce the isolation procedure was not “highly specific” (Théry et al., 2018), we cannot exclude contamination of the pellet with protein:RNA and HDL:RNA complexes. Because the *APOE* genotype could be a factor, HDL:R NA complexes likely have an impact on the final recovery of ncRNAs processed for sequencing. Very precise EV isolation and purification procedures (different approaches conducted in parallel) are required to properly address these issues.

## Conclusions

The results of this study demonstrate that SNORD115 and SNORD116 members of snoRNAs, a major class of ncRNAs, are differentially enriched in EV isolated from plasma of AD patients compared to NC. Since in our study the expression of brain specific SNORD115 cluster seems to be strongly correlated wit h AD, confirming a signature profile consisting of SNORD115 and SNORD116 in EV from plasma, postmortem brain tissue and matching plasma samples would strengthen their role as potential future biomarkers of AD. Although, the outcomes of much larger independent validation studies are difficult to predict, the results presented here point, at least, to a newly discovered association of *APOE* genotype, increased transcriptional activity of imprinted control regions, facilitated release of SNORDs in plasma EV and AD pathogenesis.

## Data Availability

The sequencing datasets are assembled in the required format and upon the acceptance of the manuscript for publication will be submitted and will be available from NCBI GEO.

## Acknowledgements

The assistance and help of: Dr. J. Milosevic in isolating RNA and preparing libraries for sequencing, of Richard Biedrzycki in processing RNAseq datasets and Dr. Florent Letronne in setting up ddPCR assays is highly acknowledged.

## List of Abbreviations

(AD): Alzheimer’s disease
(Aβ): Amyloid beta
(APOE): Apolipoprotein E
(AUC): Area Under the Curve
(SNORDs): Box C/D small nucleolar RNAs
(SNORAs): Box H/ACA small nucleolar RNAs
(CSF): Cerebrospinal fluid
(CDR): Clinical Dementia Rating
(C): Controls
(ddPCR): Droplet digital PCR
(ESCRT): Endosomal sorting complex required for transport
(exRNAs): Extracellular RNAs
(EV): Extracellular vesicles
FDR: (False Discovery Rate)
(HSP70): Heat shock protein 70
(HDL): High density lipoprotein
(miRNAs): Micro RNAs
(MMSE): Mini Mental Score Examination
(NTA): Nanoparticle Tracking Analysis
(ncRNAs): Noncoding RNAs
(PMBC): Peripheral Mononuclear Blood Cells
(PWS): Prader-Willi syndrome
(piRNAs): Piwi-interacting RNAs
(ROC): Receiver Operating Characteristic
(5-HT2C): Serotonin receptor subtype 5
(snoRNAs): Small nucleolar RNAs
(tRNAs): Transfer RNAs
(TSG101): Tumor susceptibility gene 101

## Declarations

### Ethics approval and consent to participate

All samples (Table 1 & 2) were identified through the University of Pittsburgh Alzheimer’s Disease Research Center (ADRC). The samples were collected according to protocols approved by the University of Pittsburgh Institutional Review Board and Committee for Oversight of Research Involving the Dead (CORID, No 731).

### Consent for publication: NA

### Authors’ contributions

IL, NFF and RK designed the study, analyzed the data and were major contributors in writing the manuscript. IL, RK, NFF and JW contributed to analysis of RNAseq and ddPCR datasets and data visualization. FL, NFF and KNN performed EV isolation, PCR and ddPCR data generation. MIK participated in collecting samples and APOE genotyping. All authors read and approved the manuscript.

### Authors’ information: NA

## REFERENCES

Ashton NJ, Leuzy A, Karikari TK, Mattsson-Carlgren N, Dodich A, Boccardi M, et al. The validation status of blood biomarkers of amyloid and phospho-tau assessed with the 5-phase development framework for AD biomarkers. European Journal of Nuclear Medicine and Molecular Imaging 2021.10.1007/s00259-021-05253-y

Baker EK, Butler MG, Hartin SN, Ling L, Bui M, Francis D, et al. Relationships between UBE3A and SNORD116 expression and features of autism in chromosome 15 imprinting disorders. Transl Psychiat 2020; 10(1).10.1038/s41398-020-01034-7

Bortolin-Cavaille ML, Cavaille J. The SNORD115 (H/MBII-52) and SNORD116 (H/MBII-85) gene clusters at the imprinted Prader-Willi locus generate canonical box C/D snoRNAs. Nucleic Acids Research 2012; 40(14): 6800–7.10.1093/nar/gks321

Bouchard-Bourelle P, Desjardins-Henri C, Mathurin-St-Pierre D, Deschamps-Francoeur G, Fafard-Couture É, Garant J-M, et al. snoDB: an interactive database of human snoRNA sequences, abundance and interactions. Nucleic Acids Research 2020; 48(D1): D220–D5.10.1093/nar/gkz884

Bratkovič T, Božič J, Rogelj B. Functional diversity of small nucleolar RNAs. Nucleic Acids Research 2019.10.1093/nar/gkz1140

Bratkovič T, Modic M, Camargo Ortega G, Drukker M, Rogelj B. Neuronal differentiation induces SNORD115 expression and is accompanied by post-transcriptional changes of serotonin receptor 2c mRNA. Sci Rep 2018; 8(1): 5101.10.1038/s41598-018-23293-7

Budelier MM, Bateman RJ. Biomarkers of Alzheimer Disease. J Appl Lab Med 2020; 5(1): 194–208.10.1373/jalm.2019.030080

Cassidy SB, Schwartz S, Miller JL, Driscoll DJ. Prader-Willi syndrome. Genetics in Medicine 2012; 14(1): 10–26.10.1038/gim.0b013e31822bead0

Cavaillé J. Box C/D small nucleolar RNA genes and the Prader-Willi syndrome: a complex interplay. Wiley Interdisciplinary Reviews: RNA 2017; 8(4): e1417.10.1002/wrna.1417

Cavaille J, Buiting K, Kiefmann M, Lalande M, Brannan CI, Horsthemke B, et al. Identification of brain-specific and imprinted small nucleolar RNA genes exhibiting an unusual genomic organization. Proceedings of the National Academy of Sciences 2000; 97(26): 14311–6.10.1073/pnas.250426397

Cavaillé J, Seitz H, Paulsen M, Ferguson-Smith AC, Bachellerie J-P. Identification of tandemly-repeated C/D snoRNA genes at the imprinted human 14q32 domain reminiscent of those at the Prader-Willi/Angelman syndrome region. Hum Mol Genet 2002; 11(13): 1527–38.10.1093/hmg/11.13.1527 PMID - 12045206

Chamberlain SJ. RNAs of the human chromosome 15q11-q13 imprinted region. Wiley Interdisciplinary Reviews: RNA 2013; 4(2): 155–66.10.1002/wrna.1150 PMID - 23208756

Chamberlain SJ, Lalande M. Angelman syndrome, a genomic imprinting disorder of the brain. J Neurosci 2010a; 30(30): 9958–63.10.1523/JNEUROSCI.1728-10.2010

Chamberlain SJ, Lalande M. Neurodevelopmental disorders involving genomic imprinting at human chromosome 15q11–q13. Neurobiology of Disease 2010b; 39(1): 13–20.10.1016/j.nbd.2010.03.011

Chen Y, Li G, Liu ML. Microvesicles as Emerging Biomarkers and Therapeutic Targets in Cardiometabolic Diseases. Genomics Proteomics Bioinformatics 2018; 16(1): 50–62.10.1016/j.gpb.2017.03.006

Chim SSC, Shing TKF, Hung ECW, Leung T-Y, Lau T-K, Chiu RWK, et al. Detection and Characterization of Placental MicroRNAs in Maternal Plasma. Clinical Chemistry 2008; 54(3): 482–90.10.1373/clinchem.2007.097972

Chivet M, Hemming F, Pernet-Gallay K, Fraboulet S, Sadoul R. Emerging Role of Neuronal Exosomes in the Central Nervous System. Front Physiol 2012; 3.10.3389/fphys.2012.00145

Chung MS, Langouët M, Chamberlain SJ, Carmichael GG. Prader-Willi syndrome: reflections on seminal studies and future therapies. Open Biology 2020; 10(9): 200195.10.1098/rsob.200195

Cohn WE. 5-Ribosyl uracil, a carbon-carbon ribofuranosyl nucleoside in ribonucleic acids. Biochimica et Biophysica Acta 1959; 32: 569–71.10.1016/0006-3002(59)90644-4

Deogharia M, Majumder M. Guide snoRNAs: Drivers or Passengers in Human Disease? Biology 2018; 8(1): 1.10.3390/biology8010001

Dieci G, Preti M, Montanini B. Eukaryotic snoRNAs: A paradigm for gene expression flexibility. Genomics 2009; 94(2): 83–8.10.1016/j.ygeno.2009.05.002

Driedonks TAP, Grein SGvd, Ariyurek Y, Buermans HPJ, Jekel H, Chow FWN, et al. Immune stimuli shape the small non-coding transcriptome of extracellular vesicles released by dendritic cells. Cell Mol Life Sci Cmls 2018; 75(20): 3857–75.10.1007/s00018-018-2842-8 PMID - 29808415

Dsouza VL, Adiga D, Sriharikrishnaa S, Suresh PS, Chatterjee A, Kabekkodu SP. Small nucleolar RNA and its potential role in breast cancer – A comprehensive review. Biochimica et Biophysica Acta (BBA) - Reviews on Cancer 2021; 1875(1): 188501.10.1016/j.bbcan.2020.188501

Dubois B, Feldman HH, Jacova C, Hampel H, Molinuevo JL, Blennow K, et al. Advancing research diagnostic criteria for Alzheimer’s disease: the IWG-2 criteria. The Lancet Neurology 2014; 13(6): 614–29.10.1016/s1474-4422(14)70090-0

Gao L, Ma J, Mannoor K, Guarnera MA, Shetty A, Zhan M, et al. Genome-wide small nucleolar RNA expression analysis of lung cancer by next-generation deep sequencing. Int J Cancer 2014; 136(6): E623–9.10.1002/ijc.29169 PMID - 25159866

Gstir R, Schafferer S, Scheideler M, Misslinger M, Griehl M, Daschil N, et al. Generation of a neuro-specific microarray reveals novel differentially expressed noncoding RNAs in mouse models for neurodegenerative diseases. Rna 2014; 20(12): 1929–43.10.1261/rna.047225.114

Hebras J, Marty V, Personnaz J, Mercier P, Krogh N, Nielsen H, et al. Reassessment of the involvement of Snord115 in the serotonin 2c receptor pathway in a genetically relevant mouse model. eLife 2020; 9: e60862.10.7554/eLife.60862

Irimie A, Zimta A-A, Ciocan C, Mehterov N, Dudea D, Braicu C, et al. The Unforeseen Non-Coding RNAs in Head and Neck Cancer. Genes-basel 2018; 9(3): 134.10.3390/genes9030134 PMID - 29494516

Jain G, Stuendl A, Rao P, Berulava T, Pena Centeno T, Kaurani L, et al. A combined miRNA-piRNA signature to detect Alzheimer’s disease. Transl Psychiatry 2019; 9(1): 250.10.1038/s41398-019-0579-2

Jan A, Malik MA, Rahman S, Yeo HR, Lee EJ, Abdullah TS, et al. Perspective Insights of Exosomes in Neurodegenerative Diseases: A Critical Appraisal. Frontiers in Aging Neuroscience 2017; 9: 317.10.3389/fnagi.2017.00317 PMID - 29033828

Jha P, Agrawal R, Pathak P, Kumar A, Purkait S, Mallik S, et al. Genome-wide small noncoding RNA profiling of pediatric high-grade gliomas reveals deregulation of several miRNAs, identifies downregulation of snoRNA cluster HBII-52 and delineates H3F3A and TP53 mutant-specific miRNAs and snoRNAs: Small noncoding RNA profiles in pediatric HGG. Int J Cancer 2015; 137(10): 2343–53.10.1002/ijc.29610 PMID - 25994230

Jorjani H, Kehr S, Jedlinski DJ, Gumienny R, Hertel J, Stadler PF, et al. An updated human snoRNAome. Nucleic Acids Research 2016; 44(11): 5068–82.10.1093/nar/gkw386

Keshavarz M, Krebs-Wheaton R, Refki P, Savriama Y, Zhang Y, Guenther A, et al. Natural copy number differences of tandemly repeated small nucleolar RNAs in the Prader-Willi syndrome genomic region regulate individual behavioral responses in mammals. Biorxiv 2020: 476010.10.1101/476010

Kishore S, Khanna A, Zhang Z, Hui J, Balwierz PJ, Stefan M, et al. The snoRNA MBII-52 (SNORD 115) is processed into smaller RNAs and regulates alternative splicing. Hum Mol Genet 2010; 19(7): 1153– 64.10.1093/hmg/ddp585 PMID - 20053671

Kohn MA, Carpenter CR, Newman TB. Understanding the Direction of Bias in Studies of Diagnostic Test Accuracy. Academic Emergency Medicine 2013; 20(11): 1194–206.10.1111/acem.12255

Konstantinou K, Lewis M, Dunn KM. Agreement of self-reported items and clinically assessed nerve root involvement (or sciatica) in a primary care setting. European Spine Journal 2012; 21(11): 2306–15.10.1007/s00586-012-2398-5

Lässer C, Shelke GV, Yeri A, Kim D-K, Crescitelli R, Raimondo S, et al. Two distinct extracellular RNA signatures released by a single cell type identified by microarray and next-generation sequencing. RNA biology 2016; 14(1): 0.10.1080/15476286.2016.1249092 PMID - 27791479

Lee C, Kang EY, Gandal MJ, Eskin E, Geschwind DH. Profiling allele-specific gene expression in brains from individuals with autism spectrum disorder reveals preferential minor allele usage. Nat Neurosci 2019; 22(9): 1521–32.10.1038/s41593-019-0461-9

Lefterov I, Wolfe CM, Fitz NF, Nam KN, Letronne F, Biedrzycki RJ, et al. APOE2 orchestrated differences in transcriptomic and lipidomic profiles of postmortem AD brain. Alzheimers Res Ther 2019; 11(1): 113.10.1186/s13195-019-0558-0

Liao J, Yu L, Mei Y, Guarnera M, Shen J, Li R, et al. Small nucleolar RNA signatures as biomarkers for non-small-cell lung cancer. Mol Cancer 2010; 9(1): 198.10.1186/1476-4598-9-198 PMID - 20663213

Mandrekar JN. Receiver Operating Characteristic Curve in Diagnostic Test Assessment. Journal of Thoracic Oncology 2010; 5(9): 1315–6.10.1097/jto.0b013e3181ec173d

Mao Q, Fan L, Wang X, Lin X, Cao Y, Zheng C, et al. Transcriptome-wide piRNA profiling in human brains for aging genetic factors. Jacobs J Genet 2019; 4(1)

Maoz R, Garfinkel BP, Soreq H. Alzheimer’s Disease and ncRNAs. Adv Exp Med Biol 2017; 978: 337–61.10.1007/978-3-319-53889-1_18

Milà-Alomà M, Suárez-Calvet M, Molinuevo JL. Latest advances in cerebrospinal fluid and blood biomarkers of Alzheimer’s disease. Therapeutic Advances in Neurological Disorders 2019; 12: 175628641988881.10.1177/1756286419888819

Mourksi N-E-H, Morin C, Fenouil T, Diaz J-J, Marcel V. snoRNAs Offer Novel Insight and Promising Perspectives for Lung Cancer Understanding and Management. Cells 2020; 9(3): 541.10.3390/cells9030541 PMID - 32111002

Murillo OD, Thistlethwaite W, Rozowsky J, Subramanian SL, Lucero R, Shah N, et al. exRNA Atlas Analysis Reveals Distinct Extracellular RNA Cargo Types and Their Carriers Present across Human Biofluids. Cell 2019; 177(2): 463-77.e15.10.1016/j.cell.2019.02.018

Nicholls RD, Gottlieb W, Russell LB, Davda M, Horsthemke B, Rinchik EM. Evaluation of potential models for imprinted and nonimprinted components of human chromosome 15q11-q13 syndromes by fine-structure homology mapping in the mouse. Proceedings of the National Academy of Sciences 1993; 90(5): 2050–4.10.1073/pnas.90.5.2050

Nossent AY, Ektefaie N, Wojta J, Eichelberger B, Kopp C, Panzer S, et al. Plasma Levels of snoRNAs are Associated with Platelet Activation in Patients with Peripheral Artery Disease. International Journal of Molecular Sciences 2019; 20(23): 5975.10.3390/ijms20235975 PMID - 31783567

Paolicelli RC, Bergamini G, Rajendran L. Cell-to-cell Communication by Extracellular Vesicles: Focus on Microglia. Neuroscience 2019; 405: 148–57.10.1016/j.neuroscience.2018.04.003

Pritchard AL, Harris J, Pritchard CW, Coates J, Haque S, Holder R, et al. Role of 5HT2A and 5HT2C polymorphisms in behavioural and psychological symptoms of Alzheimer’s disease. Neurobiology of Aging 2008; 29(3): 341–7.10.1016/j.neurobiolaging.2006.10.011 PMID - 17098333

Qiu W, Guo X, Lin X, Yang Q, Zhang W, Zhang Y, et al. Transcriptome-wide piRNA profiling in human brains of Alzheimer’s disease. Neurobiol Aging 2017; 57: 170–7.10.1016/j.neurobiolaging.2017.05.020

Raabe CA, Voss R, Kummerfeld D-M, Brosius J, Galiveti CR, Wolters A, et al. Ectopic expression of Snord115 in choroid plexus interferes with editing but not splicing of 5-Ht2c receptor pre-mRNA in mice. Scientific Reports 2019; 9(1): 4300.10.1038/s41598-019-39940-6 PMID - 30862860

Rajendran L, Bali J, Barr MM, Court FA, Kramer-Albers EM, Picou F, et al. Emerging roles of extracellular vesicles in the nervous system. J Neurosci 2014; 34(46): 15482–9.10.1523/JNEUROSCI.3258-14.2014

Rajendran L, Honsho M, Zahn TR, Keller P, Geiger KD, Verkade P, et al. Alzheimer’s disease beta-amyloid peptides are released in association with exosomes. Proceedings of the National Academy of Sciences 2006; 103(30): 11172–7.10.1073/pnas.0603838103

Rogelj B. Brain-Specific Small Nucleolar RNAs. Journal of Molecular Neuroscience 2006; 28(2): 103–10.10.1385/jmn:28:2:103

Ronchetti D, Mosca L, Cutrona G, Tuana G, Gentile M, Fabris S, et al. Small nucleolar RNAs as new biomarkers in chronic lymphocytic leukemia. Bmc Med Genomics 2013; 6(1): 27.10.1186/1755-8794-6-27 PMID - 24004562

Ronchetti D, Todoerti K, Tuana G, Agnelli L, Mosca L, Lionetti M, et al. The expression pattern of small nucleolar and small Cajal body-specific RNAs characterizes distinct molecular subtypes of multiple myeloma. Blood Cancer J 2012; 2(11): e96.10.1038/bcj.2012.41 PMID - 23178508

Roy J, Sarkar A, Parida S, Ghosh Z, Mallick B. Small RNA sequencing revealed dysregulated piRNAs in Alzheimer’s disease and their probable role in pathogenesis. Molecular BioSystems 2017; 13(3): 565–76.10.1039/C6MB00699J PMID - 28127595

Rozowsky J, Kitchen RR, Park JJ, Galeev TR, Diao J, Warrell J, et al. exceRpt: A Comprehensive Analytic Platform for Extracellular RNA Profiling. Cell Systems 2019; 8(4): 352-7.e3.10.1016/j.cels.2019.03.004

Runte M, Hüttenhofer A, Groß S, Kiefmann M, Horsthemke B, Buiting K. The IC-SNURF-SNRPN transcript serves as a host for multiple small nucleolar RNA species and as an antisense RNA for UBE3A. Hum Mol Genet 2001; 10(23): 2687–700.10.1093/hmg/10.23.2687 PMID - 11726556

Rynkeviciene R, Simiene J, Strainiene E, Stankevicius V, Usinskiene J, Kaubriene EM, et al. Non-Coding RNAs in Glioma. Cancers 2018; 11(1): 17.10.3390/cancers11010017 PMID - 30583549

Shaw LM, Arias J, Blennow K, Galasko D, Molinuevo JL, Salloway S, et al. Appropriate use criteria for lumbar puncture and cerebrospinal fluid testing in the diagnosis of Alzheimer’s disease. Alzheimer’s Dementia 2018; 14(11): 1505–21.10.1016/j.jalz.2018.07.220

Steinbusch MMF, Fang Y, Milner PI, Clegg PD, Young DA, Welting TJM, et al. Serum snoRNAs as biomarkers for joint ageing and post traumatic osteoarthritis. Scientific Reports 2017; 7(1): 43558.10.1038/srep43558 PMID - 28252005

Thery C, Boussac M, Veron P, Ricciardi-Castagnoli P, Raposo G, Garin J, et al. Proteomic analysis of dendritic cell-derived exosomes: a secreted subcellular compartment distinct from apoptotic vesicles. J Immunol 2001; 166(12): 7309–18.10.4049/jimmunol.166.12.7309

Théry C, Witwer KW, Aikawa E, Alcaraz MJ, Anderson JD, Andriantsitohaina R, et al. Minimal information for studies of extracellular vesicles 2018 (MISEV2018): a position statement of the International Society for Extracellular Vesicles and update of the MISEV2014 guidelines. J Extracell Vesicles 2018; 7(1): 1535750.10.1080/20013078.2018.1535750

Tosar JP, Witwer K, Cayota A. Revisiting Extracellular RNA Release, Processing, and Function. Trends in Biochemical Sciences 2021.10.1016/j.tibs.2020.12.008

Valadi H, Ekström K, Bossios A, Sjöstrand M, Lee JJ, Lötvall JO. Exosome-mediated transfer of mRNAs and microRNAs is a novel mechanism of genetic exchange between cells. Nature Cell Biology 2007; 9(6): 654–9.10.1038/ncb1596

Vendramini E, Giordan M, Giarin E, Michielotto B, Fazio G, Cazzaniga G, et al. High expression of miR-125b-2 and SNORD116 noncoding RNA clusters characterize ERG-related B cell precursor acute lymphoblastic leukemia. Oncotarget 2014; 5(0): 42398–413.10.18632/oncotarget.16392 PMID - 28415578

Wakisaka KT, Imai Y. The dawn of pirna research in various neuronal disorders. Front Biosci (Landmark Ed) 2019; 24: 1440–51

Wevrick R, Kerns JA, Francke U. Identification of a novel paternally expressed gene in the Prader-Willi syndrome region. Hum Mol Genet 1994; 3(10): 1877–82.10.1093/hmg/3.10.1877

Witwer KW. Circulating MicroRNA Biomarker Studies: Pitfalls and Potential Solutions. Clinical Chemistry 2015; 61(1): 56–63.10.1373/clinchem.2014.221341

Wright C, Shin JH, Rajpurohit A, Deep-Soboslay A, Collado-Torres L, Brandon NJ, et al. Altered expression of histamine signaling genes in autism spectrum disorder. Transl Psychiat 2017; 7(5): e1126–e.10.1038/tp.2017.87 PMID - 28485729

Xu X, Lai Y, Hua ZC. Apoptosis and apoptotic body: disease message and therapeutic target potentials. Biosci Rep 2019; 39(1).10.1042/BSR20180992

Yanfang Zhao YZ, Lei Zhang, Yanhan Dong, Hongfang Ji, Liang Shen. The Potential Markers of Circulating microRNAs and long non-coding RNAs in Alzheimer's Disease. Aging and disease 2019; 10(6): 1293–301.10.14336/ad.2018.1105

Yang H, Jiang Z, Wang S, Zhao Y, Song X, Xiao Y, et al. Long non-coding small nucleolar RNA host genes in digestive cancers. Cancer Medicine 2019; 8(18): 7693–704.10.1002/cam4.2622

Yu YT, Meier UT. RNA-guided isomerization of uridine to pseudouridine--pseudouridylation. RNA Biol 2014; 11(12): 1483–94.10.4161/15476286.2014.972855

