## Supplemental Figure 1 and 2 for "Small nucleolar RNAs in plasma and their discriminatory power as diagnostic biomarkers of Alzheimer’s disease"

### Supplemental tables and figures

#### Supplemental Figure 1.

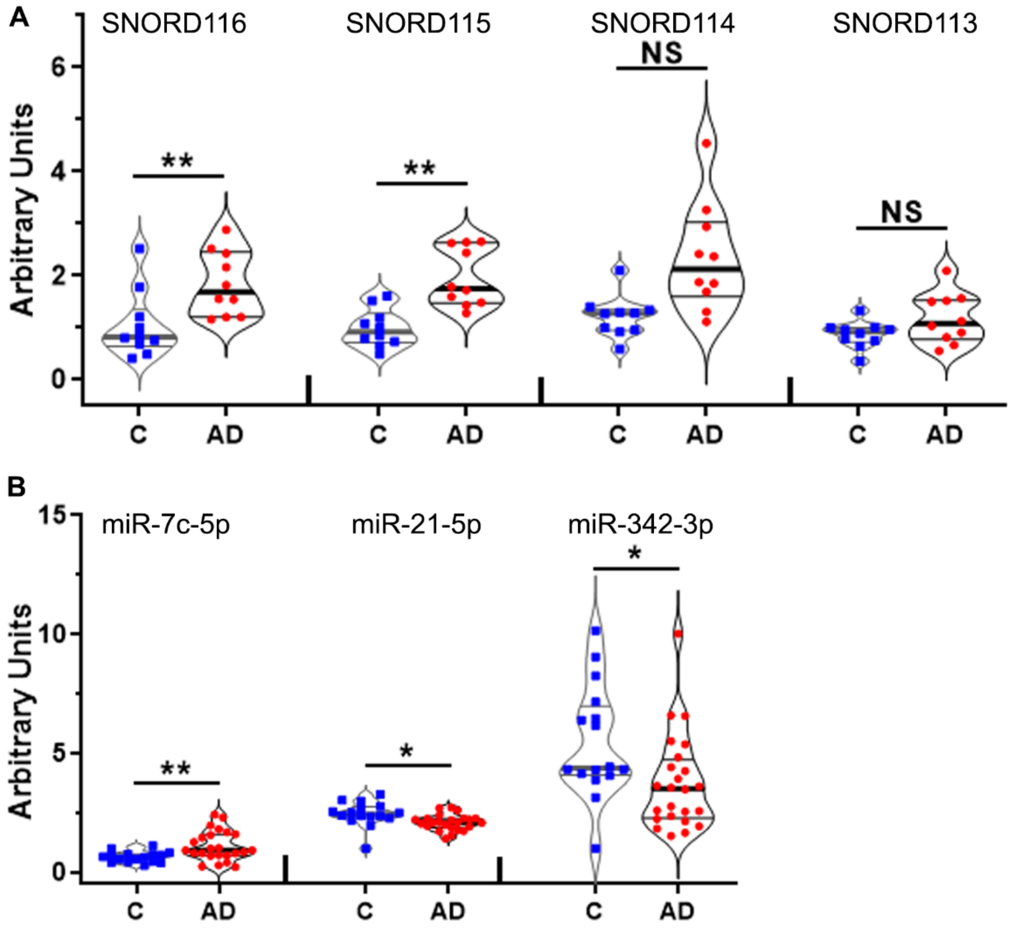


**Supplemental Figure 1. qPCR validation of RNA sequencing data for SNORDs and miRNAs; Discovery phase.** RNA isolated for RNA-seq during the Discovery phase was used for validation of select noncoding RNAs using qPCR. **A & B:** Violin plots depicting the expression of SNORDs (A; N=10 NC and AD) and miRNAs (B; N=16 NC and N=24 AD) in plasma EV from NC and AD patients. Statistical significance was determined by unpaired *t*-test. Violin plots represent kernel densities for each dataset showing median (middle line) and 75% (top) and 25% percentile (bottom). * $p < 0.05$; ** $p < 0.01$; NS = No Significance.

### Supplemental Figure 2

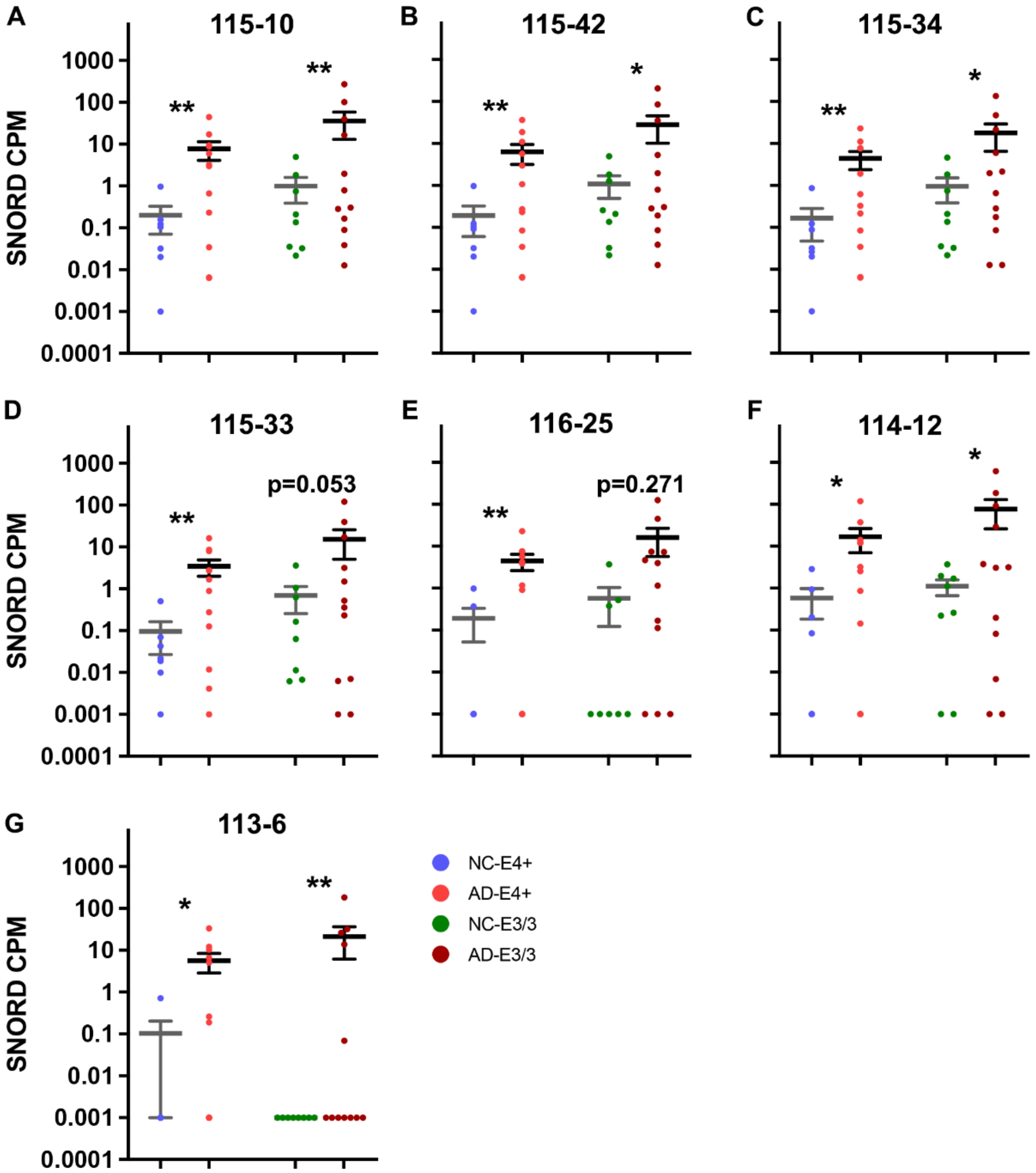


**Supplemental Figure 2. APOE isoform-dependent effect on enrichment of SNORDs in EVs of AD and NC plasma samples.** Scatter dot plots depict CPM of select SNORDs (**A-G**) from RNA-seq of plasma EVs from NC C-E3/3 (N=8), C-E4+ (N=7), AD-E3/3 (N=12) and AD-E4+ (N=12). Statistical significance was determined by exceRpt. Error bars represent mean ± SEM. ** $p<0.01$; * $p<0.05$
